# Topological relationships between perivascular spaces and progression of white matter hyperintensities: a pilot study in a sample of the Lothian Birth Cohort 1936

**DOI:** 10.1101/2021.09.18.21263770

**Authors:** Abbie Barnes, Lucia Ballerini, Maria del C. Valdés Hernández, Francesca M. Chappell, Susana Muñoz Maniega, Rozanna Meijboom, Ellen V. Backhouse, Michael S. Stringer, Roberto Duarte Coello, Rosalind Brown, Mark E. Bastin, Simon R. Cox, Ian J. Deary, Joanna M. Wardlaw

## Abstract

Enlarged perivascular spaces (PVS) and white matter hyperintensities (WMH) are features of cerebral small vessel disease which can be seen in brain magnetic resonance imaging (MRI). Given the associations and proposed mechanistic link between PVS and WMH, they are hypothesised to also have topological proximity. However, this, and the influence of their spatial proximity on WMH progression are unknown. We analysed longitudinal MRI data from 29/32 participants (mean age at baseline = 71.9 years) in a longitudinal study of cognitive ageing, from three waves of data collection at 3-year intervals, alongside semi-automatic segmentation masks for PVS and WMH, to assess relationships. The majority of deep WMH clusters were found adjacent to or enclosing PVS (Wave – 1: 77%; 2: 76%; 3: 69%), especially in frontal, parietal and temporal regions. Of the WMH clusters in the deep white matter that increased between waves, most increased around PVS (Waves – 1-2: 73%; 2-3: 72%). Formal statistical comparisons of severity of each if these two SVD markers yielded no associations between deep WMH progression and PVS proximity. These findings may suggest deep WMH clusters preferentially form and grow around PVS, possibly reflecting the consequences of impaired interstitial fluid drainage via PVS. The utility of these relationships as predictors of WMH progression remains unclear.

## Introduction

Enlarged perivascular spaces (PVS) and white matter hyperintensities (WMH) are two of the most common neuroradiological signatures of cerebral small vessel disease (cSVD) (Wardlaw *et al*., 2013b) in older age, a condition resulting from pathological processes affecting the small arteries, veins and capillaries in the brain (Shi & Wardlaw, 2016). cSVD is identified by the coexistence of microhaemorrhages, WMH, and/or lacunes/lacunar infarcts, usually accompanied by enlarged PVS, in brain magnetic resonance imaging (MRI) scans. PVS are fluid-filled spaces that surround small penetrating blood vessels (Wardlaw et al., 2013b), which provide a route for the clearance of brain waste products (Iliff et al., 2012). When enlarged, they become visible in brain MRI with the appearance of small linear or round structures depending on how they are positioned (i.e., parallel or perpendicular) with respect to the imaging plane. The cause of PVS enlargement is not fully understood but is thought to be related to impaired fluid drainage (Brown et al., 2018), as a consequence of several pathological processes that affect the cerebral microvasculature, including blood-brain barrier dysfunction, vessel stiffening, and reduced vessel pulsatility (Wardlaw, 2010; Kress et al., 2014). MRI-visible PVS, although seen also in scans of healthy adults without cSVD, have been associated with age, vascular risk factors, especially hypertension (Debette et al., 2019; Francis et al., 2019), and WMH (Wardlaw et al., 2013b). WMH, on the contrary, are considered neuropathological features and appear in a wide spectrum of disorders. They have traditionally been associated with de/dysmyelination processes and axonal degeneration (Sarbu et al., 2016), but advances in MRI have revealed that their presence may also reflect changes in interstitial fluid flow and increased water content in the white matter, especially in earlier stages of cSVD (Wardlaw et al., 2015). Like PVS, our understanding of WMH pathogenesis is poor, but they are strongly associated with vascular risk factors (Dufouil et al., 2001; Ferguson et al., 2003), other imaging features of cSVD (Ghaznawi et al., 2019), cognitive decline, gait disturbance and an increased risk of stroke and dementia (Debette & Markus, 2010).

Although PVS and WMH are commonly seen together in brain MRI scans of cognitively normal older individuals, they are clinically silent and only a few studies have analysed their relationship with conflicting results. Thus far, these studies have based their analyses on PVS and WMH severity, as determined by visual rating scores, for which subjectivity may have contributed to the conflicting findings. For instance, two cross-sectional studies found an association between the severity of enlarged PVS with WMH in two different clinical groups: lacunar stroke patients (Doubal *et al*., 2010) and community-dwelling septuagenarian individuals (Aribisala et al., 2014), in models that accounted for vascular risk factors; yet a recent meta-analysis including these studies and six others found no statistically significant associations between the two imaging markers considering results reportedly having been obtained after adjusting for vascular risk factors (Francis *et al*., 2019).

Computational methods have been developed to quantify PVS from MRI scans, helping to overcome the subjectivity associated with these scoring systems. Metrics that can be derived include total count and total volume per subject as well as length, width and volume per individual PVS (Ballerini *et al*., 2018; Boespflug *et al*., 2018). A recent study validating their use found stronger positive associations between computationally derived PVS metrics and WMH severity than visual scores (Ballerini *et al*., 2020). These cross-sectional associations suggest that widening of PVS might reflect small vessel endothelial dysfunction and impaired interstitial fluid drainage that contributes to a greater WMH burden and accumulating brain damage in ageing and cSVD (Ballerini *et al*., 2020). Longitudinal associations between these PVS metrics and WMH have not been studied, however high burden of PVS in the basal ganglia (determined by visual rating) have been associated with WMH progression after adjusting for age, sex at birth and vascular risk factors (Loos *et al*., 2015).

There is growing interest in topological relationships between PVS and WMH. Understanding how PVS and WMH are spatially related to one another in the brain may reveal important insights into their underlying mechanisms and formation. It has been observed that WMH appear to form around PVS in stroke patients (Wardlaw *et al*., 2020), typically in the parietal and posterior and lateral temporal regions. A recent study (Huang *et al*., 2021) explored the topological associations between WMH and PVS in randomly selected WMH clusters identified in a cross-sectional sample of 136 adults without previous history of stroke, brain trauma or any neurological or systemic disease, and reported that most of the randomly selected deep WMH clusters analysed were spatially connected to PVS. But longitudinal data are required to directly characterise the within-person temporal dynamics of the processes related to the evolution of WMH and PVS and their possible synergy. The findings from Huang *et al*. (2021), if confirmed in a longitudinal and more heterogeneous sample in terms of vascular disease, would provide further evidence to support a mechanistic link between both cSVD features, and reveal whether PVS could predict WMH progression. Being able to identify patients with WMH that are more likely to progress may help to prevent development of associated neurological symptoms and conditions through earlier clinical intervention. We hypothesise that in cognitively normal older adults: 1) deep WMH would more likely be spatially close to PVS and increase in size around them, and 2) those WMH clusters close to PVS, with time, will increase in size more than the WMH clusters that are distant from PVS.

## Materials and Methods

### Subjects, clinical data and imaging acquisition

We utilised brain MRI, clinical and demographic data from a randomly-selected sample of participants of the Lothian Birth Cohort 1936 Study, a longitudinal study of cognitive ageing comprising community-dwelling individuals from in and around Edinburgh born in 1936. All participants provided written consent to take part in the study under protocols approved by Lothian (REC 07/MRE00/58) and Scottish Multicentre (MREC/01/0/56) Research Ethics Committees. The methods used for MRI acquisition and clinical data in this cohort have been reported previously (Deary *et al*., 2007; Wardlaw *et al*., 2011; Taylor *et al*., 2018). Participants had their first brain MRI scan at the second wave of testing at mean age of 72.6 years, and subsequent MRI examinations spaced at 3-year intervals (Taylor *et al*., 2018). For this study we randomly (i.e., using a random numbers generator function in MATLAB R2019b) selected a sample of 32 participants with brain MRI available for the first 3 consecutive scanning waves (i.e., a total of 96 individual MRI scans). All brain images were acquired at the Western General Hospital of Edinburgh in a GE Signa Horizon HDx 1.5 T clinical scanner (General Electric, Milwaukee, WI) following the research acquisition protocol described in Wardlaw *et al*. (2011). Vascular risk factors, which included presence or absence of hypertension, hypercholesterolaemia, diabetes mellitus, stroke and history of cardiovascular disease, were self-reported at each wave (Taylor *et al*., 2018).

### Image Processing

We used existent binary masks of WMH from the three waves and PVS in the centrum semiovale from the first imaging wave generated as previously published (Valdés Hernández et al., 2010, 2020; Ballerini et al., 2020). Briefly, for the first scanning wave (i.e., referred hereby as wave 1) WMH masks were generated using a multispectral method that combines T2*-weighted and fluid attenuated inversion recovery (FLAIR) images mapped in the red-green colour space and quantised to facilitate a robust thresholding using minimum variance quantisation (Valdés Hernández et al., 2010). For the other two waves WMH binary masks were generated from a statistical analysis of the FLAIR-normalised intensities, seeking full compatibility with the semi-automatic approach applied in the first wave, and higher level of automation. All WMH masks were checked and manually edited to ensure the segmentation was accurate. Segmentation masks for PVS in the deep white matter (i.e., excluding the internal and external capsules that are part of the region clinically defined as basal ganglia for purposes of PVS identification) were generated using a computational method described previously (Ballerini et al., 2016, 2020), on T2-weighted images. Succinctly, tubular structures are enhanced using the Frangi filter, optimised for this purpose with the parameters described by Ballerini et al. (2016). The output from the filter is thresholded, binarised and quantified in the region of interest. Manually rectifying PVS segmentation masks was not feasible, however the images were checked for noise and other artefacts that would affect accuracy of PVS segmentation.

### Visual assessments

Adjacency/closeness (or not) of PVS with WMH clusters was recorded while/by visually inspecting all slices where PVS were segmented. Initially, the inspection was done in axial slices, and double-checked in all radiological orientations throughout the sample in all waves. All visual assessments, performed with MRIcron v1.0.20190902 (https://www.nitrc.org/projects/mricron/), were blinded to visual rating scores for PVS and WMH, and participants’ clinical and demographic information. These were repeated by the same observer to ensure perfect intra-observer agreement (mean differences in PVS count ± 95% confidence interval are equal to −0.207 ± 1.637 for Wave 1 and - 0.034 ± 1.361 for Wave 2, see Bland-Altman plot in Supplementary Figure 1). Deep WMH clusters were defined as regions or voxel clusters of WMH not contiguous with the WMH located in the periventricular lining. Periventricular WMH caps surrounding the horns of the lateral ventricles were counted as deep WMH clusters if they extend more than 3 mm into the deep white matter (Kim, Macfall and Payne, 2008). Cross-sectional topological relationships were assessed on the first scanning wave. Each deep WMH cluster was recorded and classified as either “close” or “not close” to a PVS in the first scan in co-registered T2-weighted and FLAIR images after superimposing the PVS and WMH binary masks in different contrasting colours (Figure 1). Deep WMH were defined as ‘close’ to a PVS if their segmentation was overlapping or contiguous with a PVS and ‘not close’ if it was not overlapping or contiguous. Adjacent slices were carefully inspected to ensure not to miss or double-count any PVS. Each deep WMH cluster was also labelled and the region of the brain it was found in was recorded - either frontal, parietal, temporal or occipital, determined by examining lobar segmentations from a digital anatomic atlas (Debowski, 2018).

**Figure 1.**
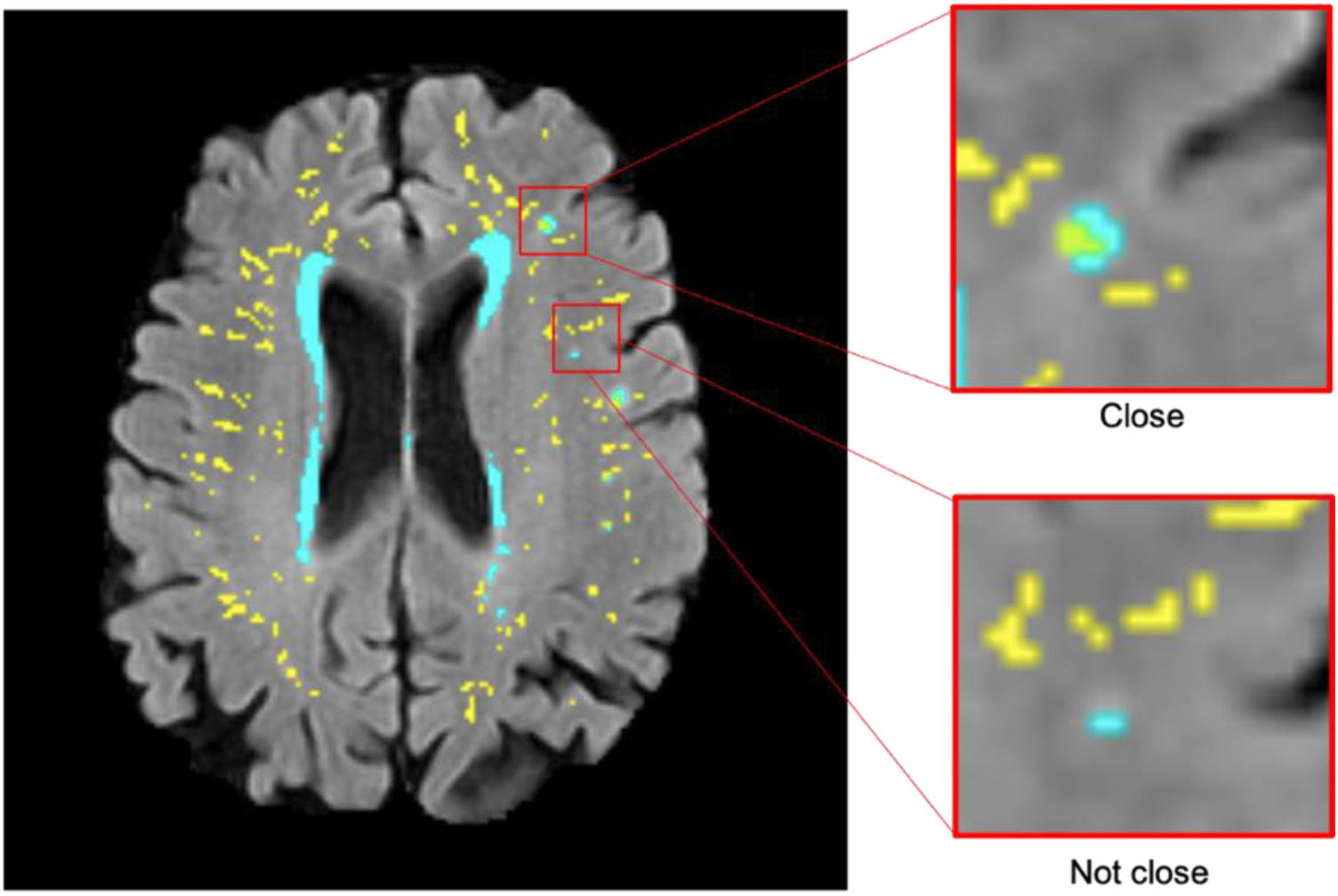
Examples of deep WMH in the baseline scan that would be classified as ‘close’ and ‘not close’ to baseline PVS when determining cross-sectional topological relationships (segmentation masks for WMH in cyan and PVS in yellow, overlaid on FLAIR MRI).

To identify topological relationships longitudinally, the total number of deep WMH clusters were counted again. This time, each was classified according to how their morphology changed relative to baseline PVS between consecutive waves (e.g., waves 1 and 2 and waves 2 and 3) in each participant. A further mask that represented the change in WMH between waves was also overlaid in the FLAIR image to facilitate this assessment (Figure 2). These relationships were defined as follows: ‘increase around’ – deep WMH cluster increased so that more voxels of the WMH mask became contiguous with a PVS; ‘increase close’ – deep WMH cluster previously considered ‘close’ to a PVS, increased in size, but no more of it became contiguous with a PVS (i.e., the adjacency between both – PVS and WMH – masks observed was owed to the same number of voxels as in the previous wave); ‘increase not close’ – deep WMH cluster increased but it was still not contiguous with a PVS; ‘no increase’ – no visible change in the deep WMH cluster (Figure 2). Again, all deep WMH clusters counted were recorded according to the lobar brain region.

**Figure 2.**
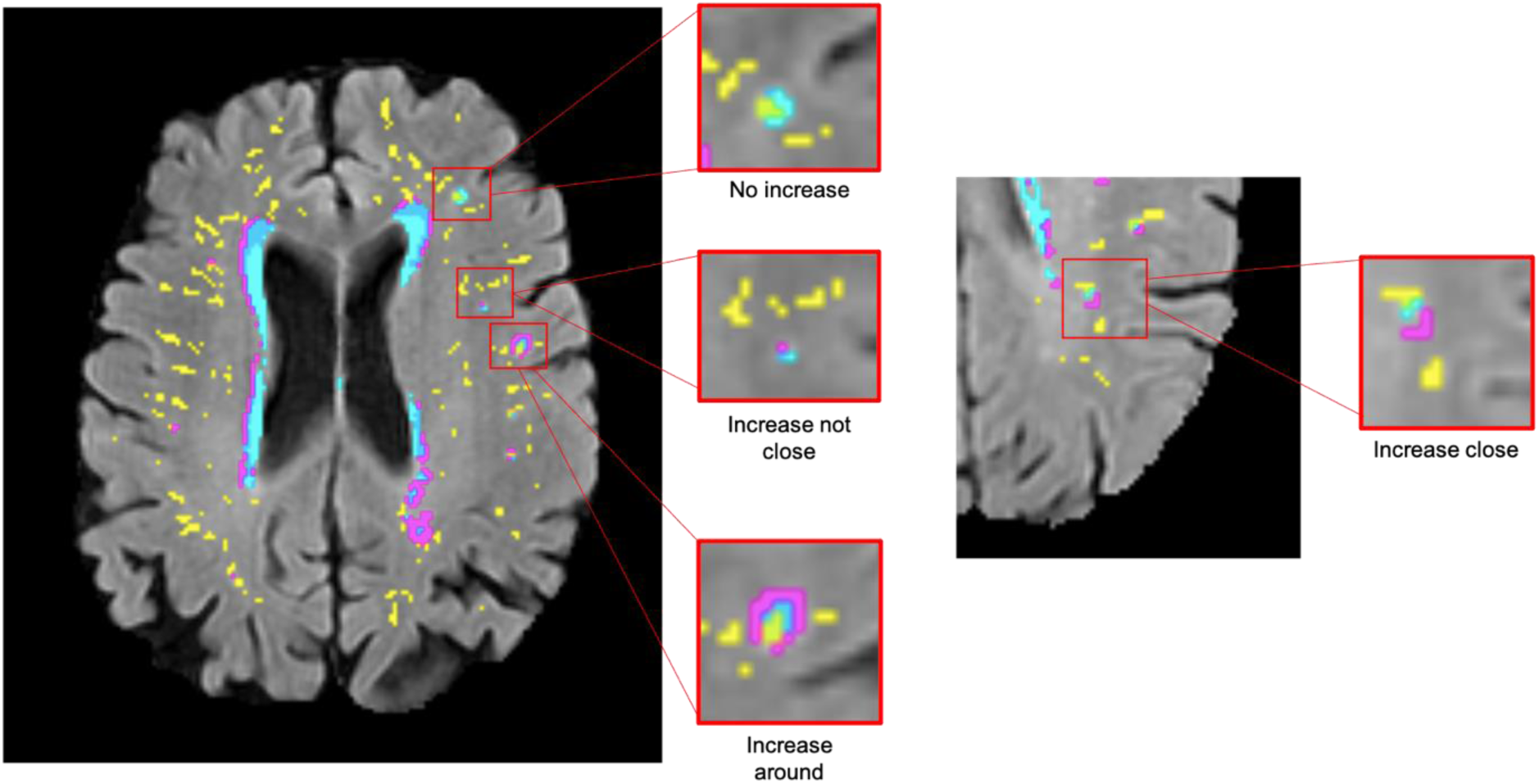
Examples of deep WMH for each of the different categories used to determine longitudinal topological relationships (segmentation masks for WMH in cyan, change in WMH in violet and PVS in yellow, overlaid on FLAIR MRI).

### Statistical analysis

Statistical analysis was performed in R (version 3.6.2). First, the counted data per overall individual brain scan was summarised to show the median count and distribution within the sample, per category. Cross-sectional and longitudinal topological relationships were analysed by calculating percentages per category, both overall and per brain region. To allow longitudinal analysis of the cross-sectional relationships, change in count of deep WMH clusters ‘close’ and ‘not close’ to baseline PVS between waves was simplified into a binary variable where ‘1’ indicated an increase in the number of deep WMH clusters and ‘0’ indicated otherwise (i.e., no change or a decrease). Binary logistic regression with random effects was used to model the association between the change in number of deep WMH clusters and their location relative to a PVS. Random effects accounted for variation between participants. Data per overall brain was used and separate models were performed for waves 1 to 2, and waves 2 to 3. As well as unadjusted models, we evaluated models that corrected for sex at birth and vascular risk factors including hypertension, hypercholesterolaemia, diabetes mellitus and history of cardiovascular disease, with a maximum of three predictors per model. We estimated our sample size guided by the only study that visually assessed the topological relationships between WMH and PVS (Huang et al. (2021), which randomly selected 600 deep WMH clusters from a cross-sectional sample for analysis. In our sample, at baseline (i.e., only in Wave 1), the total number of deep WMH clusters counted surpasses 600. Resources available for this study allowed us to process data from 29 individuals. Nevertheless, given the exploratory nature of this study, the complexity and nature of the assessments, to ensure reproducibility, comparability and objectivity in the analyses, this estimate was considered appropriate (Hoenig & Heisei, 2001).

## Results

### Sample characteristics

Data from three participants were excluded due to noise on their T2-weighted images which visually was considered to have affected PVS segmentation. One participant was not included in the models performed between waves 1 and 2 as no deep WMH cluster were counted in either of these waves. The mean age of the sample at baseline was 71.87 years (SD = 0.38). The follow-up MRI scans from waves 2 and 3 were obtained three and six years later respectively (i.e., at mean age approximately 75 and 78 years). Table 1 shows the baseline characteristics of the sample, including sex at birth, vascular risk factors, and Fazekas visual rating scores of WMH.

**Table 1.** Baseline participant characteristics, n = 29 (CVD = cardiovascular disease).

|  | Male | Female |
| --- | --- | --- |
| Gender | 17 (58.62%) | 12 (41.38%) |
|  | Deep | Periventricular |
| WMH Fazekas scores (median [QR1 QR3]) | 1 [1 1] | 1 [1 2] |
|  | Yes | No |
| Diabetes | 1 (3.45%) | 28 (96.55%) |
| Hypertension | 14 (48.28%) | 15 (51.72%) |
| Hypercholesterolaemia | 9 (31.03%) | 20 (68.97%) |
| Stroke | 0 (0.00%) | 29 (100.00%) |
| History of CVD | 9 (31.03%) | 20 (68.97%) |

### Cross-sectional topological relationships

Figure 3 shows some examples that illustrate the variations in the cross-sectional topologies observed. For deep WMH clusters classed as ‘close’ to a PVS, most bordered a PVS with a small area of overlap (Figure 3(a)), however some completely encircled a PVS, completely overlapping with it (Figure 3(b)). Some of the larger deep WMH clusters closely related to PVS were also seen to be continuous or overlapping with multiple PVS (Figure 3(c)). Within deep WMH clusters that were classed as ‘not close’ to a PVS, there was also considerable variability in the distance separating them. Some were fairly close to a PVS but not continuous with it while others were much further from a PVS (Figure 3(d)). Many of these different topologies existed simultaneously in the same participant (Figure 4).

**Figure 3.**
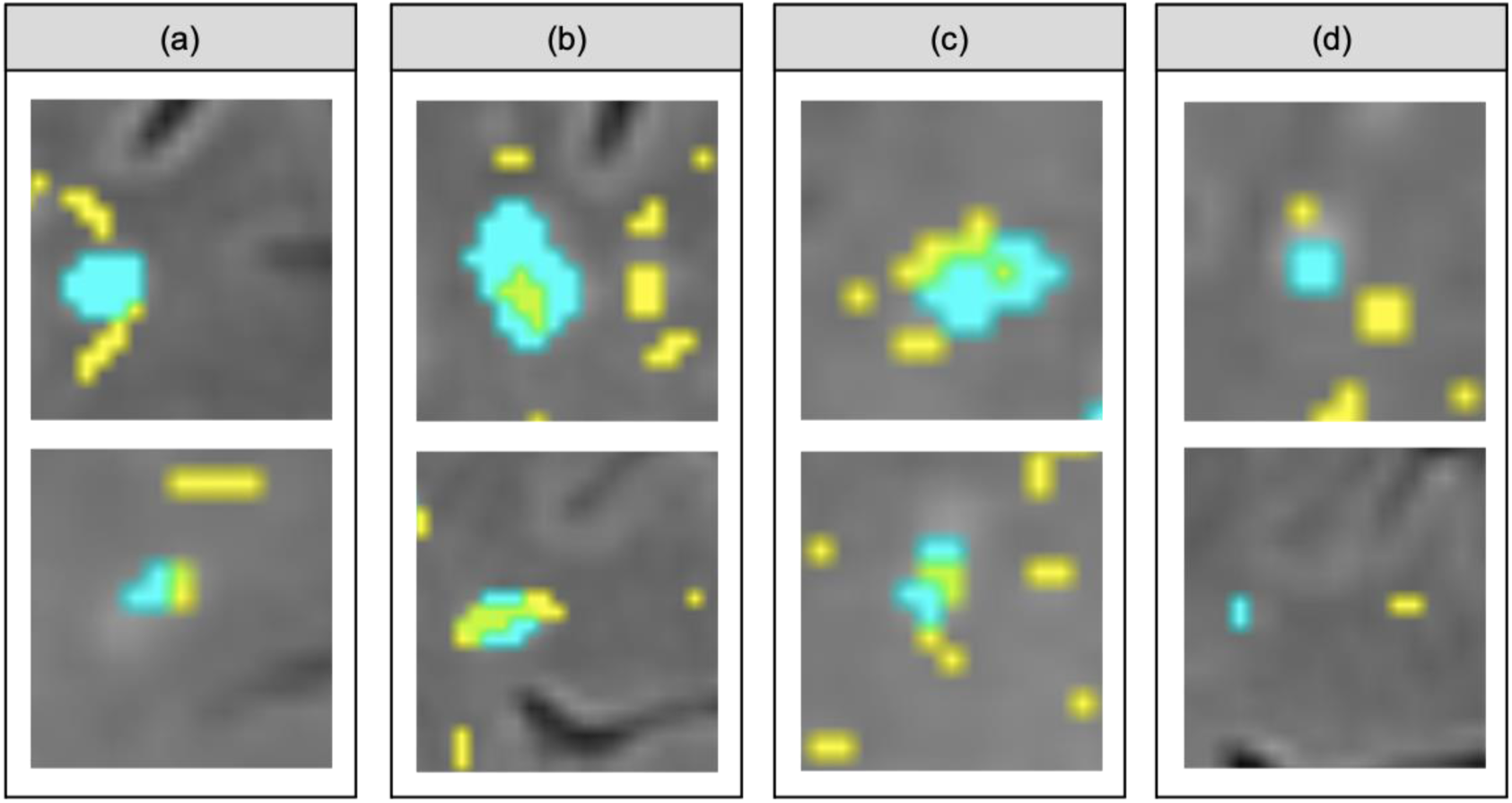
Examples of the different topological types observed within the ‘close’ vs. ‘not close’ classification used for describing cross-sectional topological relationships. (a), (b) and (c) are variations within deep WMH classed as ‘close’, while (d) are variations within those classed as ‘not close’ (segmentation masks for WMH in cyan and PVS in yellow, overlaid on FLAIR MRI).

**Figure 4.**
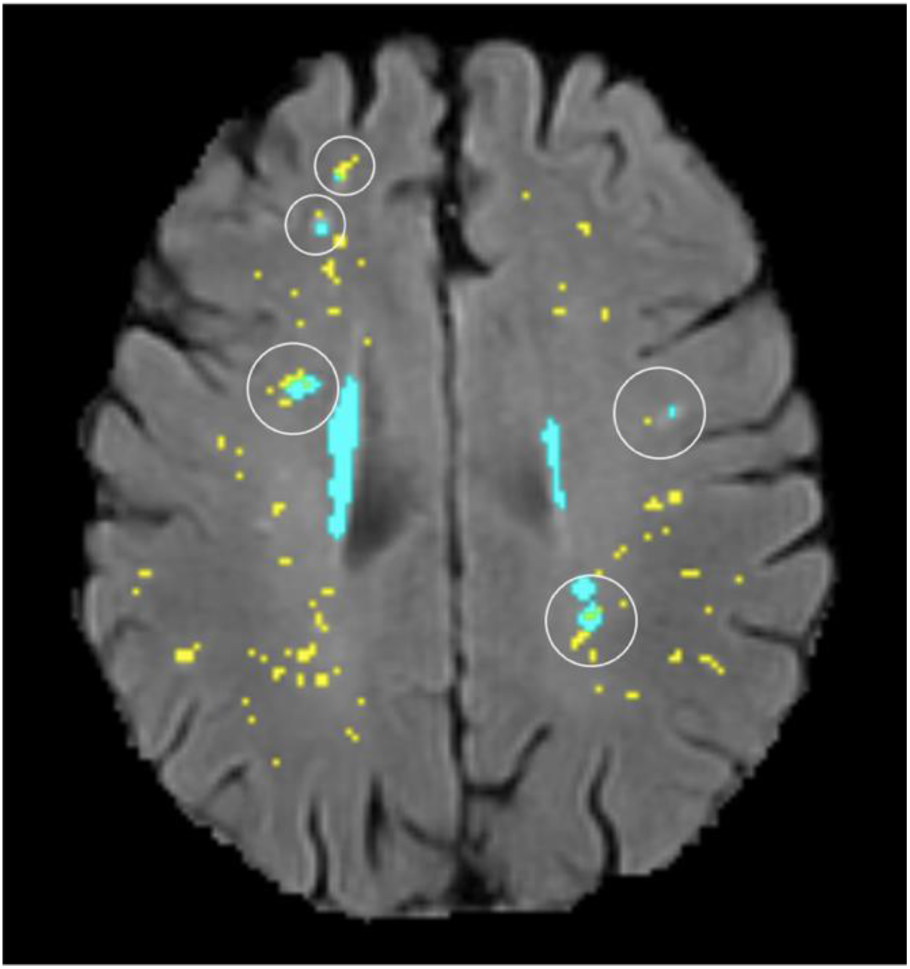
Illustration of several different topological types (circled) existing in the same participant (segmentation masks for WMH in cyan and PVS in yellow, overlaid on FLAIR MRI).

Median count of deep WMH clusters ‘close’ to baseline PVS was greater than the median count of those ‘not close’ at all time-points (Figure 5). The counts were not normally distributed in this sample, with a positive skew due to several participants having zero or very low counts of deep WMH clusters. Percentages were calculated from the total numbers of deep WMH clusters found spatially ‘close’ and ‘not close’ to baseline PVS across the sample (Table 2). A higher percentage of WMH clusters in Wave 1 were found close to PVS in this wave than not close. In the rest of the waves this pattern was also observed (Wave 1: 77% ‘close’, 23% ‘not close’; Wave 2: 76% ‘close’, 24% ‘not close’; Wave 3: n = 69% ‘close’, 31% ‘not close’) (Figure 6, Table 2). Despite far fewer deep WMH clusters being counted in the parietal and temporal regions than in the frontal region (Table 3), the percentages of these that were found ‘close’ to baseline PVS (i.e., PVS identified at the first wave) were higher than those found ‘not close’ in these regions (Figure 7), (for instance, Wave 1 – frontal: 75% ‘close’, 25% ‘not close’; parietal: 81% ‘close’, 19% ‘not close’; temporal: 86% ‘close’, 14% ‘not close’). The occipital region was not included in Figure 7 as only 1 deep WMH cluster was counted in Wave 1, skewing the percentages.

**Table 2.** Total number of deep WMH counted across the sample in waves 1, 2 and 3 that were ‘close’ and ‘not close’ to baseline (i.e., Wave 1) PVS, per overall brain.

|  | Close (%) | Not close (%) | Total |
| --- | --- | --- | --- |
| Wave 1 | 479 (76.64) | 146 (23.36) | 625 |
| Wave 2 | 585 (75.68) | 188 (24.32) | 773 |
| Wave 3 | 909 (69.12) | 406 (30.87) | 1315 |

**Table 3.** Total number of deep WMH counted across the sample in waves 1, 2 and 3 that were ‘close’ and ‘not close’ to baseline PVS, per lobar region.

|  | Close (%) | Not close (%) | Total |
| --- | --- | --- | --- |
| **Wave 1** |  |  |  |
| Frontal | 343 (54.88) | 114 (18.24) | 457 (73.12) |
| Parietal | 109 (17.44) | 26 (4.16) | 135 (21.60) |
| Temporal | 25 (4.00) | 4 (0.64) | 29 (4.64) |
| Occipital | 1 (0.16) | 0 (0.00) | 1 (0.16) |
| **Wave 2** |  |  |  |
| Frontal | 401 (51.87) | 125 (16.17) | 526 (68.05) |
| Parietal | 146 (18.89) | 42 (5.43) | 188 (24.32) |
| Temporal | 34 (4.40) | 14 (1.81) | 48 (6.21) |
| Occipital | 3 (0.39) | 5 (0.65) | 8 (1.03) |
| **Wave 3** |  |  |  |
| Frontal | 633 (48.14) | 264 (20.08) | 897 (68.21) |
| Parietal | 206 (15.66) | 74 (5.63) | 280 (21.29) |
| Temporal | 60 (4.56) | 42 (0.03) | 102 (7.76) |
| Occipital | 10 (0.76) | 22 (1.67) | 32 (2.43) |

**Figure 5.**
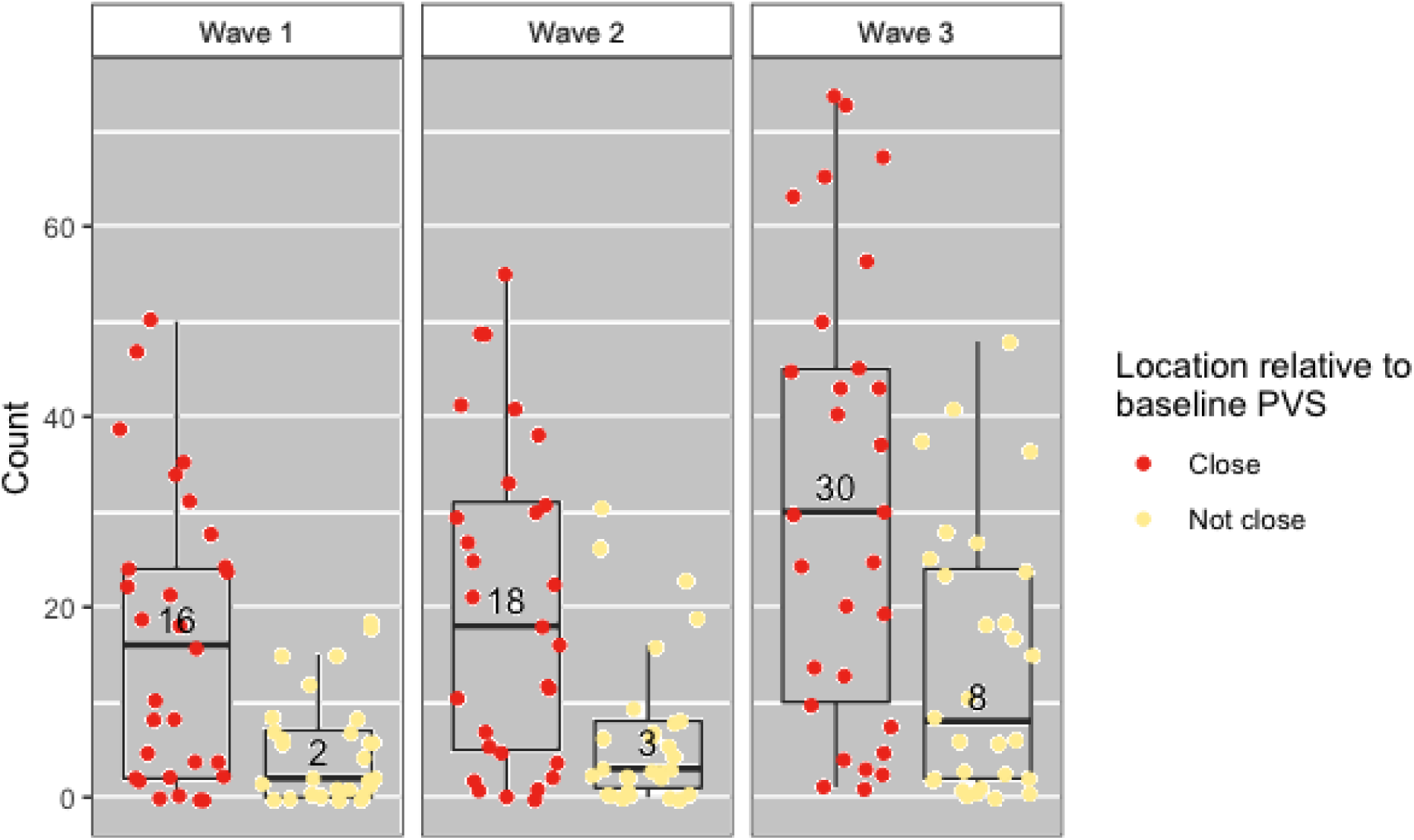
Distribution of counts of deep WMH in waves 1, 2 and 3, found close and not close to baseline (i.e. wave 1) PVS. The points plotted on each boxplot represent a different participant in the sample and the numbers inside the boxplots correspond to the median count.

**Figure 6.**
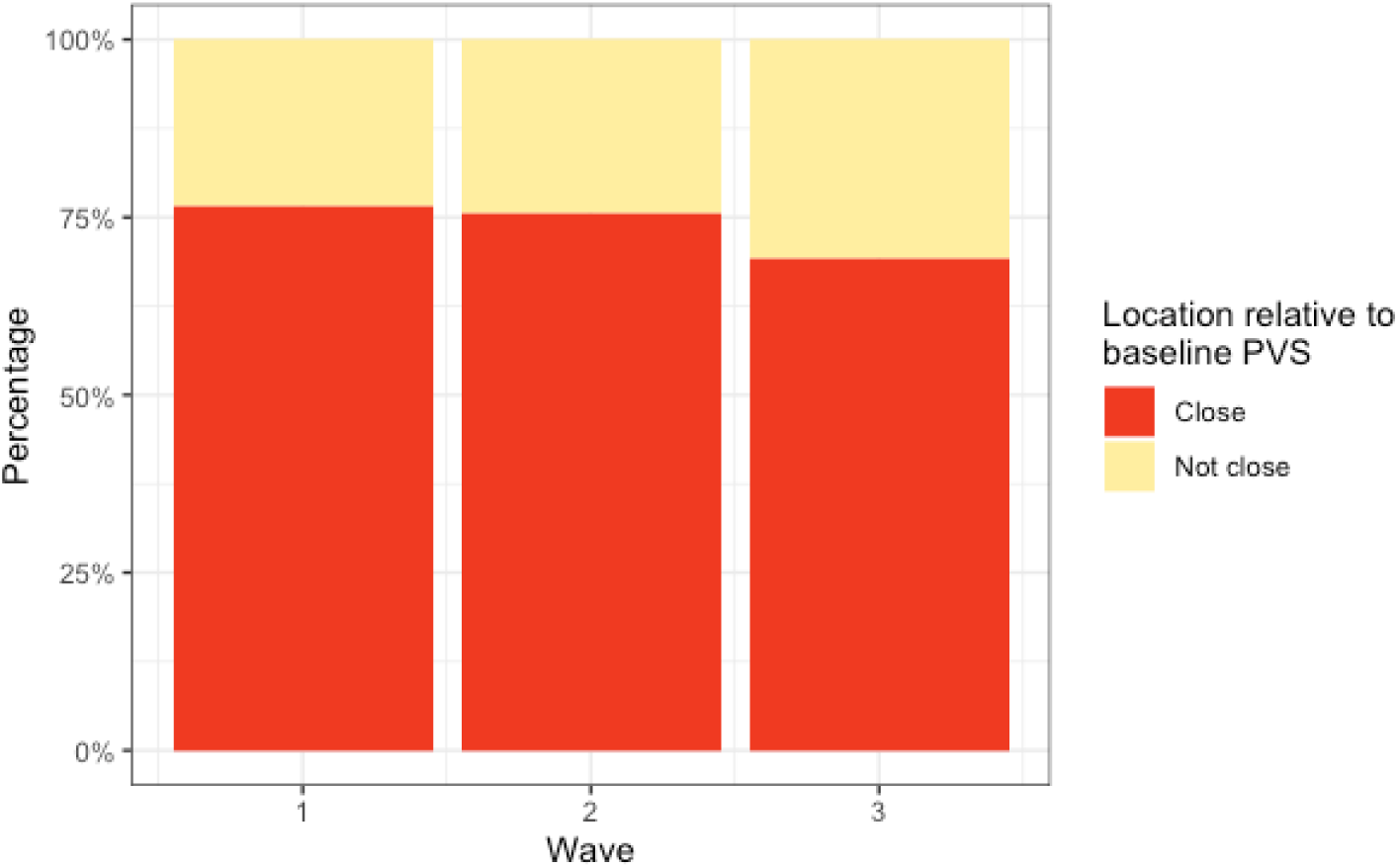
Percentages of deep WMH in waves 1, 2 and 3 that were found ‘close’ and ‘not close’ to baseline (i.e., Wave 1) PVS.

**Figure 7.**
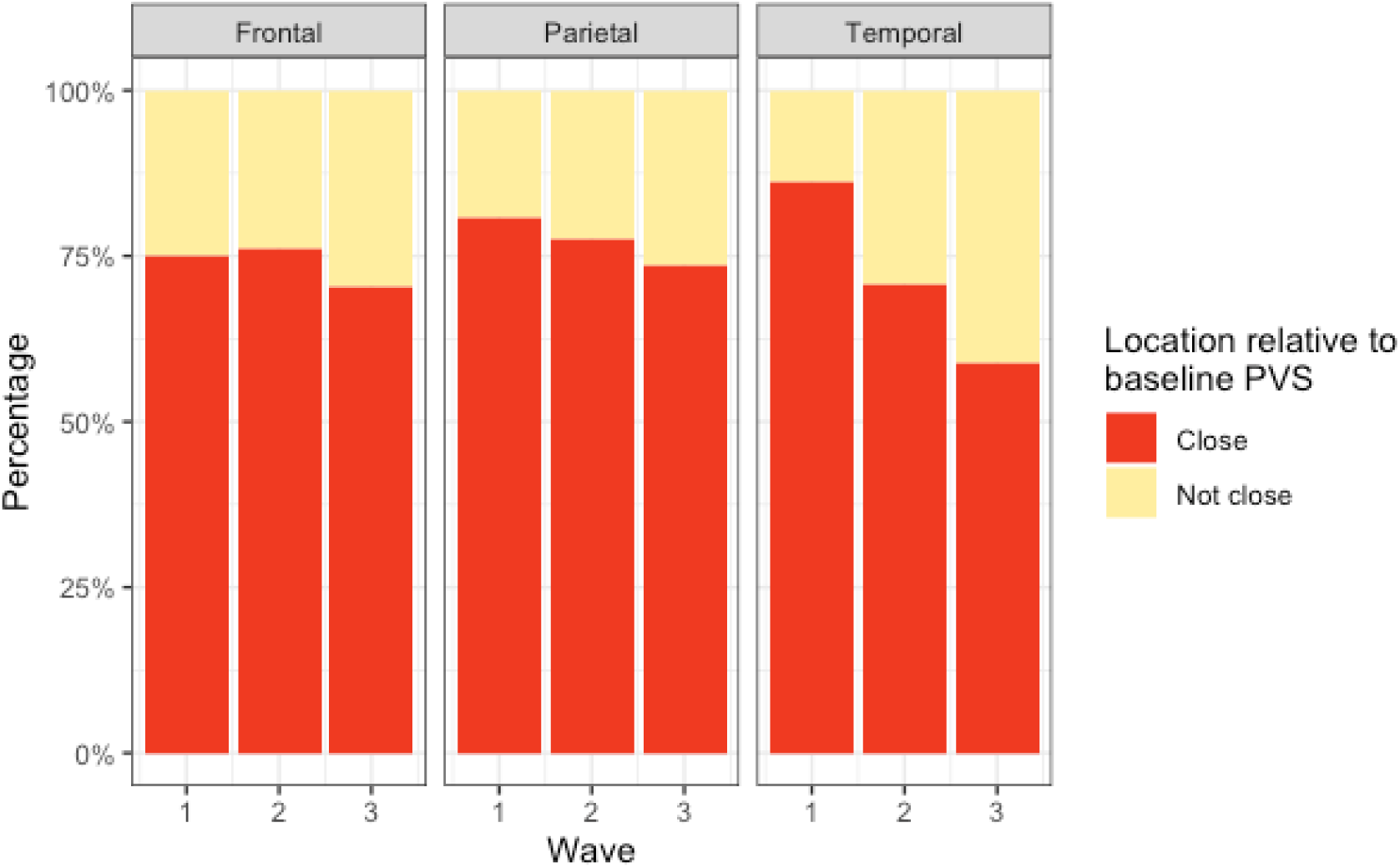
Percentages of deep WMH in waves 1, 2 and 3 that were found close and not close to baseline PVS in the frontal, parietal and temporal regions.

### Associations between progression of deep WMH clusters and proximity to PVS at Wave 1

Between waves 1 and 2, the number of deep WMH clusters changed in 28 participants. There was a median increase of 2 in deep WMH clusters spatially close to baseline PVS (i.e., PVS detected at Wave 1), and 1 in those not close. Between waves 2 and 3, the number of deep WMH clusters changed for all 29 participants. The median change in number spatially close to a PVS increased in 10, while the median change for those ‘not close’ increased in 6. Logistic regression models with random effects found no significant associations between progression (defined by an increase in number) of deep WMH clusters and proximity of these clusters to PVS (i.e., either ‘close’ or ‘not close’) at Wave 1. Table 4 shows the odds ratios, 95% confidence intervals and p values for all models performed.

**Table 4.** Association between proximity of deep WMH to PVS and deep WMH progression by binary logistic regression with random effects (OR = odds ratio, 95% CI = 95% confidence interval, VRF = vascular risk factors). OR (95% CI; p-value)

| OR (95% CI; p-value) |  |  |
| --- | --- | --- |
|  | Unadjusted | Adjusted for sex and VRF |
| **Wave 1 to 2** |  |  |
| Not close | 1.00 | 1.00 |
| Close | 1.93 (0.58-6.83; p = 0.28) | 2.44 (0.68-8.82; p = 0.17) |

| Wave 2 to 3 |  |  |
| --- | --- | --- |
| Not close | 1.00 | 1.00 |
| Close | 4.05 (0.09-176.00; p = 0.47) | 2.00 (0.16-25.10; p = 0.59) |

### Longitudinal topological relationships

The counts of changes in deep WMH clusters’ morphology in relation to baseline PVS were not normally distributed and positively skewed (Figure 8). The percentages of the different changes between waves 1 and 2 and waves 2 and 3 are displayed in Figure 9. These percentages were calculated from the total number of deep WMH clusters counted in each category in waves 1 and 2 respectively (i.e., first waves in each time interval), seen in Table 5. A greater percentage of deep WMH clusters increased in size (Wave 1 to 2: 70% increased, 30% did not increase; Wave 2 to 3: 77% increased, 23% did not increase). Of those that increased, the most frequent change was an increase in size around the location of the already existent (i.e., “baseline”) PVS (Wave 1 to 2: 73% ‘increased around’, 10% ‘increased close’, 17% ‘increased not close’; Wave 2 to 3: 72% ‘increased around’, 10% ‘increased close’, 18% ‘increased not close’). When lobar regions were analysed separately, these trends were also observed in the frontal, parietal and temporal regions, as seen in Figure 10 (Table 6). The occipital region was not included in Figure 10 because, as previously referred, only one WMH cluster was identified in this region at Wave 1.

**Table 5.** Total number of deep WMH counted across the sample that increased around, close and not close to baseline PVS and did not increase between waves 1 and 2 and waves 2 and 3, per overall brain.

|  | Increase around | Increase close | Increase not near | No increase | Total |
| --- | --- | --- | --- | --- | --- |
| Wave 1 to 2 | 319 | 45 | 76 | 189 | 629 |
| Wave 2 to 3 | 425 | 60 | 108 | 179 | 772 |

**Table 6.** Total number of deep WMH counted across the sample that increased around, close and not close to baseline PVS and number of those that did not increase between waves 1 and 2 and waves 2 and 3, per region.

|  | Increase around | Increase close | Increase not near | No increase | Total |
| --- | --- | --- | --- | --- | --- |
| **Wave 1 to 2** |  |  |  |  |  |
| Frontal | 227 | 33 | 62 | 139 | 461 |
| Parietal | 74 | 8 | 12 | 44 | 138 |
| Temporal | 17 | 4 | 2 | 6 | 29 |
| Occipital | 1 | 0 | 0 | 0 | 1 |
| **Wave 2 to 3** |  |  |  |  |  |
| Frontal | 283 | 42 | 79 | 124 | 528 |
| Parietal | 110 | 17 | 23 | 40 | 190 |
| Temporal | 29 | 1 | 4 | 12 | 46 |
| Occipital | 3 | 0 | 2 | 3 | 8 |

**Figure 8.**
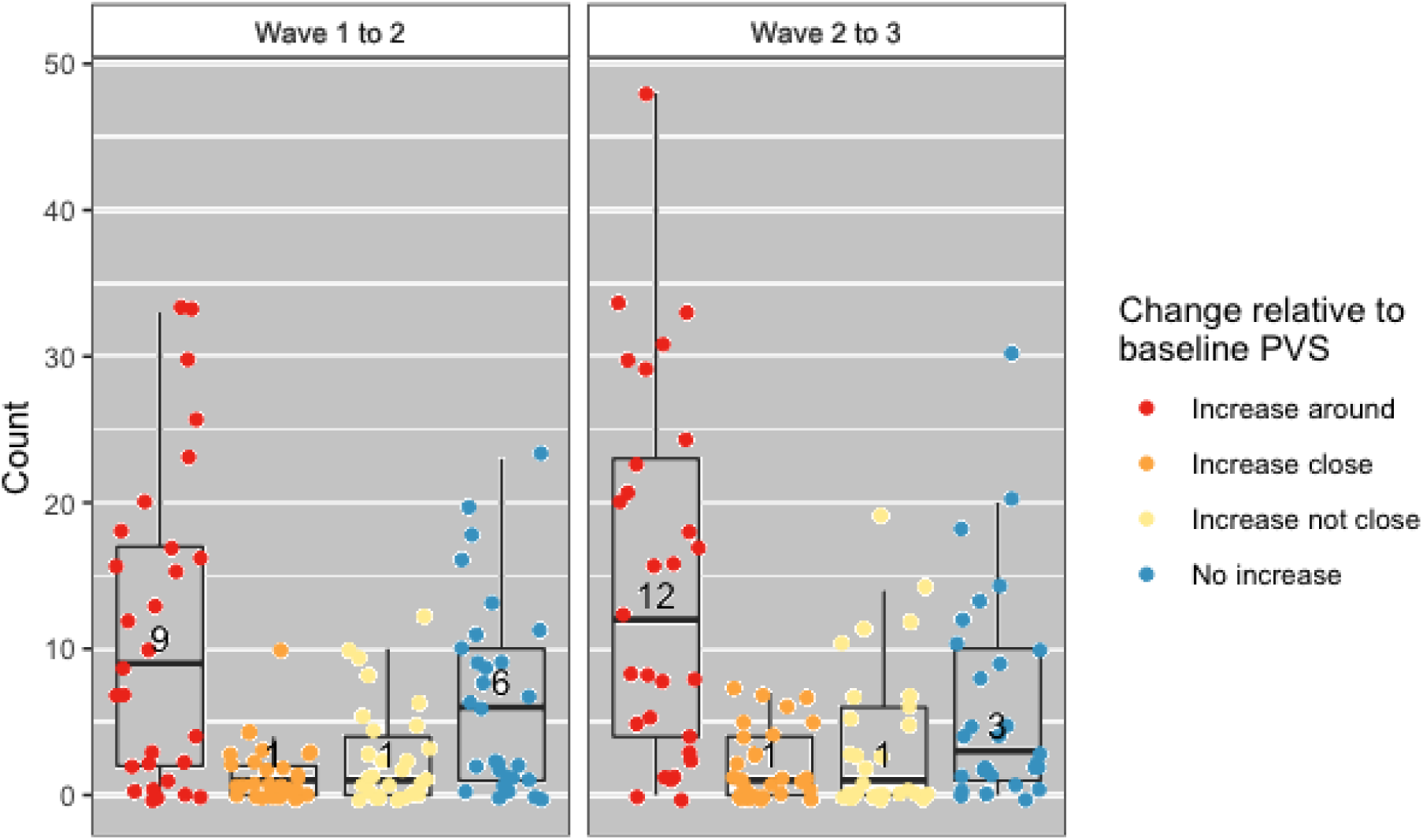
Distribution of counts of deep WMH that increased around, close and not close to baseline PVS and did not increase between waves 1 and 2 and waves 2 and 3. The points plotted on each boxplot represent a different participant in the sample and the numbers represent the median count.

**Figure 9.**
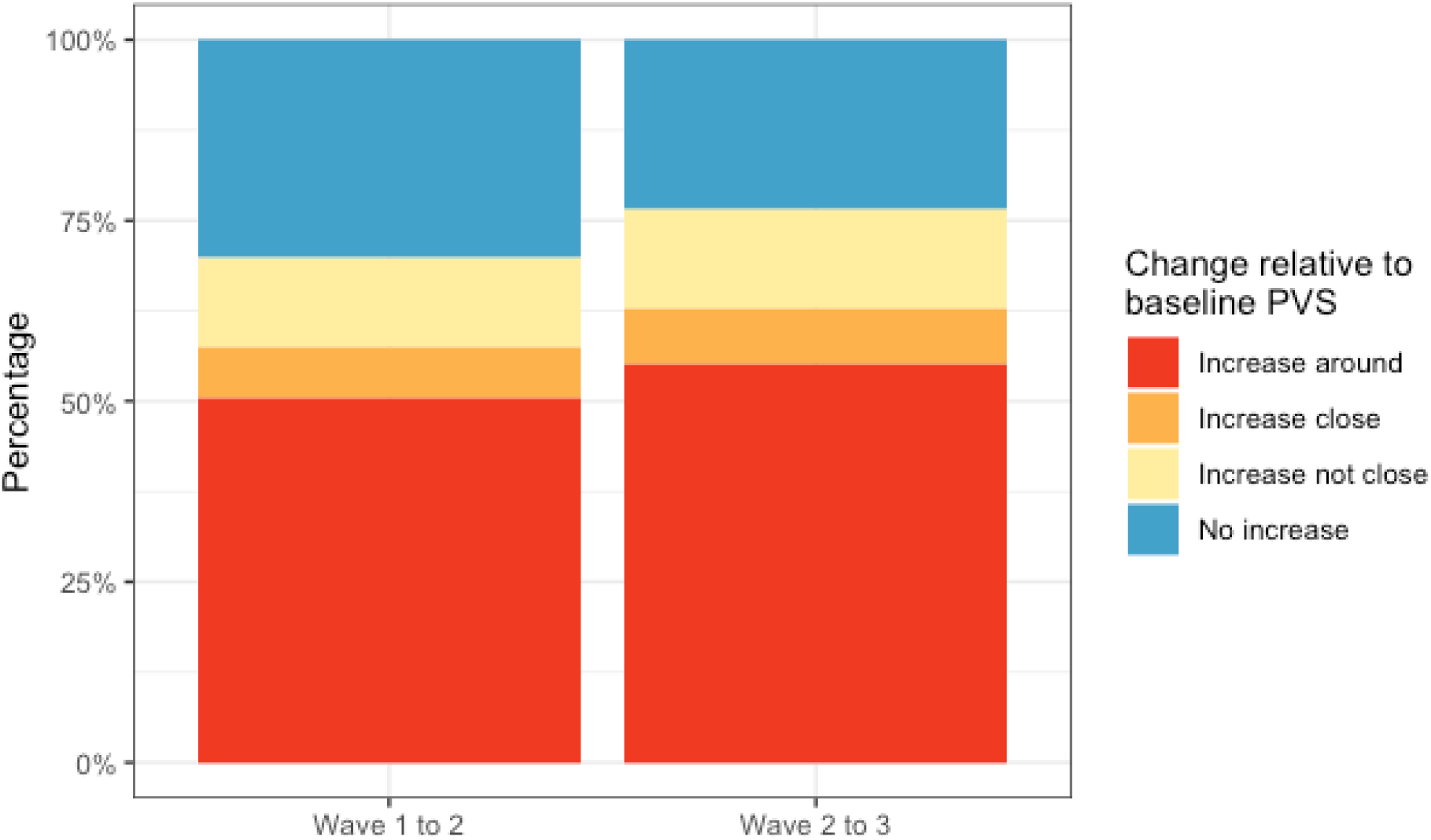
Percentages of deep WMH that increased around, close and not close to baseline PVS and did not increase between waves 1 and 2 and waves 2 and 3.

**Figure 10.**
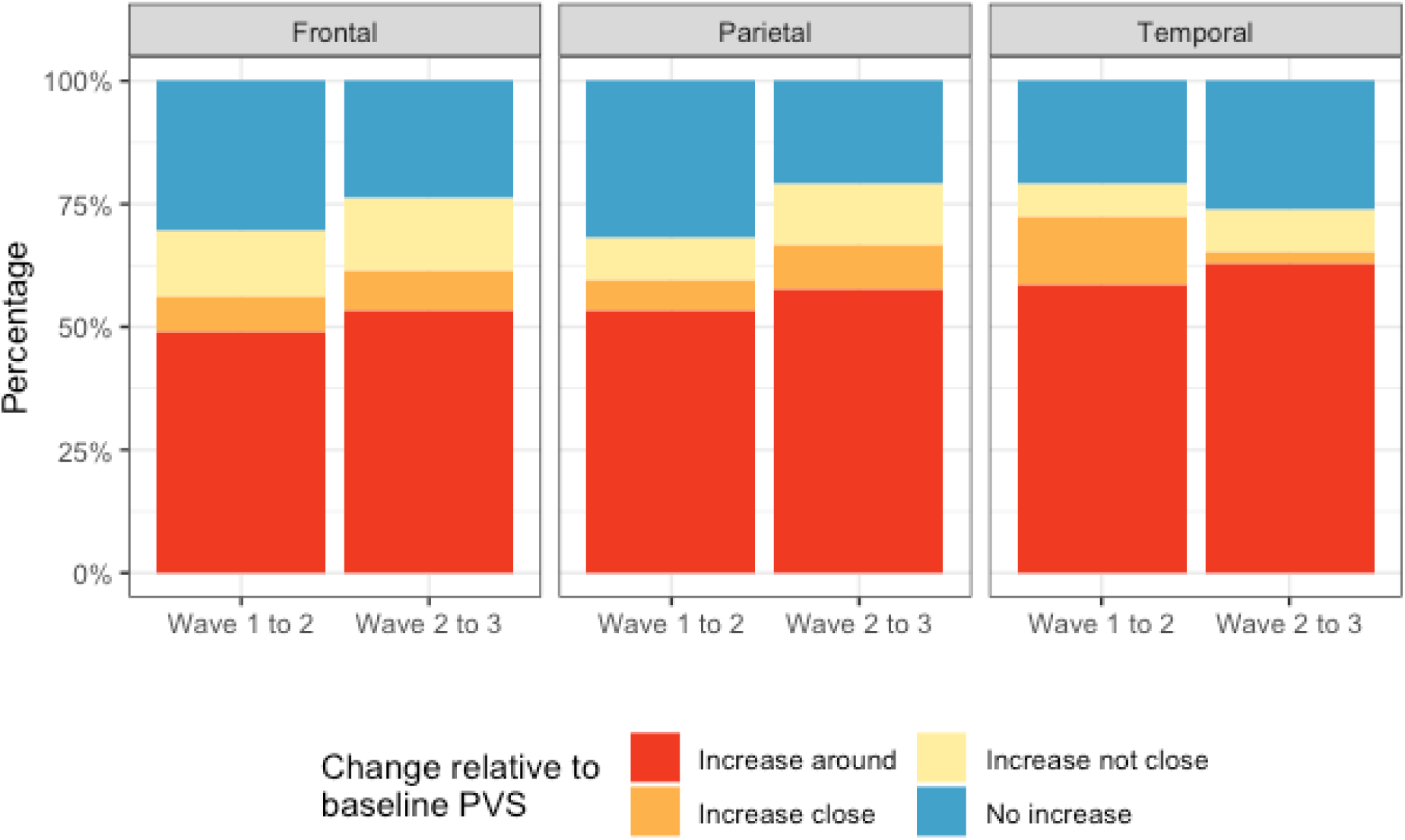
Percentages of deep WMH that increased around, close and not close to baseline PVS and did not increase in the frontal, parietal and temporal regions, between waves 1 and 2 and waves 2 and 3.

Although, also as previously referred, far fewer deep WMH clusters were counted in parietal and temporal regions compared with the frontal region, we more frequently observed larger WMH clusters forming around multiple PVS in the parietal and posterior temporal regions (Figure 11). Deep WMH clusters in the frontal region were typically smaller in volume with a small area of continuity with a PVS.

**Figure 11.**
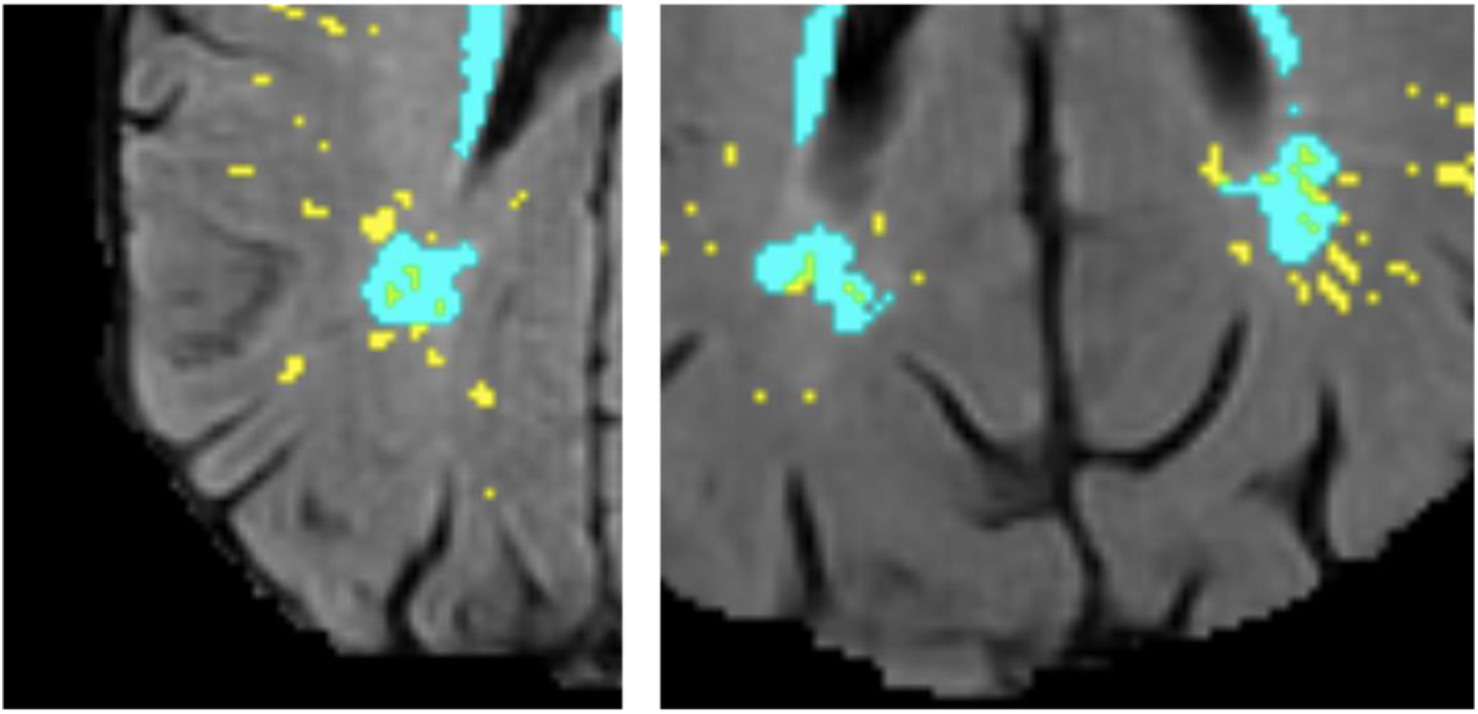
Examples of large deep WMH in the parietal region that were found spatially close to multiple PVS (segmentation masks for WMH in cyan and PVS in yellow, overlaid on FLAIR MRI).

## Discussion

This pilot study in a sample of participants from a community-dwelling Scottish cohort is the first longitudinal study to date that evaluated the topological spatial relationship between PVS and deep WMH clusters. By carefully analysing data across six years acquired in three equally-spaced time points, our study provides further insights into this novel area of research by corroborating the hypothesis that most deep WMH increase in size and form around PVS, previously inferred from cross-sectional data. Our results, therefore, suggest that in normal ageing, WMH formation may be linked to impaired brain clearance mechanisms in addition to the vascular origin referred to in the current literature (Wardlaw et al., 2013). From our observations it is possible to infer that worsening interstitial fluid drainage over time can cause further accumulation of fluid in the white matter immediately adjacent to where drainage via PVS was previously impaired. This is consistent with the proposed mediating role of free water in the brain in the association seen between PVS and deep WMH clusters by a previous cross-sectional study (Huang et al., 2021).

Clinical reviews have previously proposed that WMH may preferentially form around PVS (Wardlaw, 2010; Wardlaw *et al*., 2013a). Another review that examined the pathological evidence for the failure of the brain clearance mechanisms as a significant cause of the overall pathology found particularly in the ageing brain, stated that enlarged PVS reflect impaired interstitial fluid drainage and lead to the development of WMH (Weller et al., 2015). Our study for the first time provides evidence to support this claim, by analysing the spatial proximity, distribution and evolution of WMH in relation to PVS in brain scans from an age-homogeneous cohort representative of normal ageing across 6 years within the eight decade of life.

Previously published data about topological relationships observed a marked difference between the percentages of WMH clusters spatially connected and not connected to PVS (Huang et al., 2021) when compared to our study. However, this may be due to several differences in study design. The age of the sample utilised by Huang *et al*. (2021) was younger, with an inter-quartile range of 56-65 years, compared to our sample, where participants were aged approximately 72, 75 and 78 years when relationships were analysed. As both PVS and WMH are independently correlated with increasing age (De Leeuw *et al*., 2001; Zhu *et al*., 2011), this may have influenced the differences seen. In terms of vascular disease our sample is also more heterogeneous as it includes individuals with cardiovascular disease and total Fazekas scores ranging from one to five. Also, different from our study, Huang and colleagues analysed a random sample of deep WMH clusters rather than all present. While this was more feasible in a larger sample and allowed better appreciation of relationships in 3D, it may have introduced sampling error.

Although our longitudinal analysis revealed an increase in size of the WMH clusters close to or overlapping with the PVS identified at the first wave of scanning, our statistical analysis suggests that the number of deep WMH clusters spatially close to PVS were no more likely to increase than those spatially not close and that location of deep WMH clusters in relation to PVS at the first scan was not a predictor for WMH progression in this sample. There are several possible reasons why no statistically significant associations were found. Although deemed appropriate, it is likely that a sample size of 29 was not sufficiently large. Given the small sample, its homogeneity in terms of age compensated the likelihood of biasedness of the adjusted statistical models, as age was not a factor needed to account for. Nevertheless, the number of outcome events per predictor variable were slightly less than the recommended 10 (Peduzzi *et al*., 1996). Binary logistic regression with random effects was chosen because the alternatives were deemed beyond the scope of this study due to the complexity and number of assessments, which such a small sample size would be unlikely to support. A larger study would use more sophisticated analysis methods such as Poisson regression to fully exploit the count-nature of the data. (Coxe *et al*., 2009). Defining deep WMH clusters’ progression as an increase in their number for statistical analyses was a further limitation as it didn’t take into account change in volume of individual clusters and the possibility of multiple clusters coalescing into one over time, which was observed in some cases. For such a detailed analysis to be performed computationally it would have required access to a large “ground truth” databank currently non-existent. Also, the analysis of the odds of deep WMH clusters close to PVS increasing in number to those not close to PVS is complex since it is unclear what deep WMH clusters not close to PVS represent. Although their appearance is unlikely to be related to the consequences of impaired interstitial fluid drainage, they may be just as likely to increase in number for different reasons.

Another limitation of our study is the relatively low spatial resolution of the images for the assessment of these type of structures (i.e., 1 × 1 × 2 mm^3^ at 1.5 Tesla), which may have introduced an error in counting the PVS that occur parallel to the axial plane. Relying on segmentation to define the relationships seen, despite improving the reliability and reproducibility in the assessments, meant that accuracy of the data collected was dependent on the accuracy of segmentation, particularly for PVS for which is impractical to be visually checked and edited individually, especially for cases with a high burden of them. Inaccuracies in the segmentation of baseline PVS in this cohort as a whole have been recognised, including misclassification due to variations in gyral patterns (Ballerini *et al*., 2020). It is not clear whether punctate WMH that pathology studies recognise as WMH composed of and characterised by enlarged PVS (Chimowitz *et al*., 1992; Fazekas *et al*., 1993; Munoz *et al*., 1993) are segmented as PVS, WMH, or both. This may have also affected how accurately deep WMH clusters were classified, as well as the possibility that segmentation masks for PVS and WMH may have obscured one another when overlaid in MRI. Lastly, due to the visual and 3D nature of the assessments, despite the high intra-observer reliability, one cannot discard the possibility of potentially mistaken classification of WMH clusters close to PVS diagonally across slices, or double counting of the same cluster in different planes.

The fact that only one observer performed the visual assessments limited the sources of variation in the dataset. Counting all deep WMH clusters is very time-consuming, especially in participants with a high WMH burden, so the methodology used here would not be feasible in large samples. Nonetheless, it proved a useful way of obtaining data about topological relationships between deep WMH clusters and PVS, both cross-sectionally and longitudinally, to test our hypotheses. Due to the novelty of this research there is no reliable annotated data to train any machine-learning algorithm to reliably classify large data fully automatically. Existent automatic classifiers that demand less or no data at all have a suboptimal accuracy for being applied in clinical studies or test clinically-relevant hypotheses. Therefore, future research should continue to use the methodology developed for this study to generate ground truth data in a larger sample, reassess the relationships seen, and re-evaluate the utility of the presented paradigm to determine its usefulness in predicting WMH progression. Moreover, future studies should include in the analyses participants with history of previous strokes to investigate differences in tendencies over time (if they exist) between individuals who had a stroke against those who did not. It will be also useful to validate the results against the percentage overlap between segmentation masks for PVS and WMH, which would better take into account differences in volume. As described in the Methods section and in previous publications from this cohort, WMH binary masks separately for periventricular and deep regions are not currently available, and the available PVS segmentations only cover the white matter in the corona radiata supraventricular of the centrum semiovale (Ballerini et al., 2020, Valdés Hernández et al., 2020). Therefore, with the data available at present such computational analysis would have been misleading.

Since our findings support the idea that PVS enlargement precedes WMH development, future research on better understanding what causes PVS enlargement would be important. MRI-visible PVS in the centrum semiovale are linked to amyloid deposition (Charidimou *et al*., 2015; Martinez-Ramirez *et al*., 2018), which increases blood brain barrier permeability (Bell *et al*., 2012), therefore ways to prevent this would be clinically beneficial.

## Conclusions

In this pilot study most deep WMH clusters were found spatially close to a baseline PVS and of those that progressed with time, most increased around a baseline PVS. Although this sample is very small, these findings support a mechanistic link between these two cSVD features that may improve our understanding of the mechanisms involved in cSVD and WMH development to help reduce and prevent associated symptoms and neurological conditions. Formal statistical comparisons of severity of these two SVD markers yielded no associations between deep WMH clusters progression and the location of these clusters relative to PVS. The sample size should be increased to confirm these associations. Future research should also explore more feasible ways of analysing these relationships (i.e., automatically) and the causes of PVS enlargement to continue furthering our understanding of the mechanisms involved in cSVD.

## Data Availability

The magnetic resonance imaging data used in this manuscript is publicly available via BRAINS Imagebank https://www.brainsimagebank.ac.uk/

## Acknowledgements

We thank all those involved with the Lothian Birth Cohort 1936 Study, including the participants, nurses, clinicians, researchers and support staff, without whom the data utilised in this project would not have been available. We especially thank Dr. David Alexander Dickie, Dr. Stewart Wiseman, and Prof. Benjamin S. Aribisala for their contributions to the development or generation of WMH segmentations throughout the different waves of the LBC 1936 Study. This study is partially funded by the Selfridges Group Foundation under the Novel Biomarkers 2019 scheme (ref UB190097) administered by the Weston Brain Institute. The LBC1936 is supported by Age UK as The Disconnected Mind Project (http://www.disconnectedmind.ed.ac.uk), the Medical Research Council [G1001245/96099] and The University of Edinburgh. LBC1936 MRI brain imaging was supported by Medical Research Council (MRC) grants [G0701120], [G1001245], [MR/M013111/1] and [MR/R024065/1]. Magnetic Resonance Image acquisition and analyses were conducted at the Brain Research Imaging Centre, Neuroimaging Sciences, University of Edinburgh (www.bric.ed.ac.uk) which is part of SINAPSE (Scottish Imaging Network—A Platform for Scientific Excellence) collaboration (www.sinapse.ac.uk) funded by the Scottish Funding Council and the Chief Scientist Office. This work was supported by the Centre for Cognitive Ageing and Cognitive Epidemiology, funded by the Medical Research Council and the Biotechnology and Biological Sciences Research Council (MR/K026992/1), the Row Fogo Charitable Trust (BRO-D.FID3668413), the European Union Horizon 2020, (PHC-03-15, project No 666881), SVDs@Target, the Fondation Leducq Transatlantic Network of Excellence for the Study of Perivascular Spaces in Small Vessel Disease, ref no. 16 CVD 05, the US National Institutes of Health (R01AG054628), a Sir Henry Dale Fellowship jointly funded by the Wellcome Trust, the Royal Society (SRC, Grant Number 221890/Z/20/Z), and the Medical Research Council UK Dementia Research Institute at the University of Edinburgh.

## Notes

### Competing Interest Statement

The authors have declared no competing interest.

### Clinical Trial

Observational cohort study on cognitive ageing and cognitive epidemiology

### Clinical Protocols

https://doi.org/10.1038/s41598-018-19781-5

https://doi.org/10.1186/1471-2318-7-28

https://doi.org/10.1093/ije/dyy022

https://doi.org/10.1007/s00330-010-1718-6

https://doi.org/10.1111/j.1747-4949.2011.00683.x

### Author Declarations

All participants provided written consent to take part in the study under protocols approved by Lothian (REC 07/MRE00/58) and Scottish Multicentre (MREC/01/0/56) Research Ethics Committees.

